# Etiologies of bloody diarrhea in children presenting with acute gastroenteritis to US emergency departments

**DOI:** 10.1101/2024.04.03.24305279

**Authors:** Paola Fonseca-Romero, Sharia M. Ahmed, Ben J. Brintz, D. Matthew Vierkant, Jennifer Dien Bard, Daniel M. Cohen, Ara Festekjian, Amy L. Leber, Jami T. Jackson, Neena Kanwar, Chari Larsen, Rangaraj Selvarangan, Kimberle C. Chapin, Andrew T. Pavia, Daniel T. Leung, IMPACT study investigators

**Affiliations:** Department of Pathology, Spencer Fox Eccles School of Medicine, University of Utah, Salt Lake City, UT, USA; Department of Internal Medicine, Spencer Fox Eccles School of Medicine, University of Utah, Salt Lake City, UT, USA; Children’s Hospital, Los Angeles; Keck School of Medicine, University of Southern California, Los Angeles, CA, US; Department of Pediatrics, Spencer Fox Eccles School of Medicine, University of Utah, Salt Lake City, UT, USA; Department of Pediatrics, Nationwide Children’s Hospital, Columbus, OH, USA; Children’s Mercy Hospital, Kansas City, MO, USA; Deepull, Barcelona, Spain; Warren Alpert Medical School, Brown University, Providence, RI, USA

**Keywords:** Gastroenteritis, bloody diarrhea, antibiotic, multiplex panel, pediatric

## Abstract

Among 111 children presenting with bloody diarrhea in a multicenter study of molecular testing in US emergency departments, we found viral pathogens in 18%, bacteria in 48%, protozoa in 2%, and no pathogens detected in 38%.

## Introduction

Diarrheal diseases continue to be the leading cause of death globally in children under five, with an estimated 1.7 billion cases and 525,000 deaths every year[1]. In the United States alone, diarrheal illness accounts for 178.8 million cases, resulting in 474,000 hospitalizations and 5,000 deaths each year[2,3]. Bloody diarrhea, or hematochezia associated with a diarrheal illness, arises from intestinal inflammation[4] and may be accompanied by dehydration, fever, and abdominal pain. Bloody diarrhea can lead to substantial healthcare utilization and medical costs [5,6]. Despite this, our knowledge of the causes of bloody diarrhea is limited mainly to single-center studies, many of which were from the pre-molecular era.

Bloody diarrhea is often associated with *Shigella spp and* other invasive enteropathogens such as *Shiga-toxin producing E. coli (STEC)*, Salmonella *spp, Campylobacter spp, and Yersinia spp*[5,7–10]. However, recent studies have shown that many cases of bloody diarrhea cannot be attributed to only bacteria, that STEC only accounts for a minority of cases, and in many cases, infectious etiology cannot be determined[4,11]. The Infectious Disease Society of America (IDSA) guidelines recommend that patients presenting with fever or bloody stool be evaluated for enteropathogens, for which antimicrobial agents may be clinically beneficial[12]. Historically, given the long turnaround time of stool culture, the use of antibiotics in cases of bloody diarrhea has been empiric. However, overuse of antimicrobials is associated with adverse medication effects and hemolytic-uremic syndrome (HUS) in STEC infection[13], and on the population level, may lead to the development of antimicrobial resistance. In one US study, 13% of children with acute gastroenteritis were prescribed antibiotics at outpatient visits [14]. Knowledge of etiologies in children with bloody diarrhea may improve the appropriate use of antibiotics and diagnostics. Therefore, our primary objective in this study was to determine the etiological agents responsible for bloody diarrhea in children.

## Methods

### Study Design and Setting

We performed a secondary analysis of data collected as part of a pragmatic stepped wedge study of the impact of multiplex PCR testing (BioFire FilmArray GI Panel, bioMerieux, Salt Lake City, Utah), which tests for 22 pathogens: 5 viruses, 13 bacteria, and 4 protozoa in five US pediatric emergency departments, also known as the Implementation of Molecular Diagnostic for Pediatric Acute Gastroenteritis study (IMPACT)[15], conducted from April 2015 to September 2016 (Supplemental Figure 1). Written informed consent was obtained from parents or legal guardians, and children provided age-appropriate assent. The trained study coordinator administered a questionnaire on symptoms, medical history, treatment, demographics, and epidemiologic exposures of children presenting with gastroenteritis. During the visit or within 48 hours, we asked all children to provide a stool sample for PCR multiplex testing.

### Eligibility

Patients eligible were < 18 years old with symptoms of gastroenteritis (diarrhea, vomiting, abdominal pain) and presented to the ED or on-site urgent care center. The IMPACT study had 1157 eligible patients (Supplemental Figure 2). Excluding 137 patients who did not have diarrhea or whose status was unknown, among the 1020 patients who reported diarrhea, we included 126 (12.3%) who reported bloody diarrhea in this analysis.

### Etiological Outcomes

We considered multiple pathogens detected in a single sample as co-infections. In this analysis, we did not consider enteropathogenic E. coli (EPEC) in children of any age and *C. difficile* in children <=2 years of age given that they are of unclear clinical significance.

### Statistical analysis

We conducted data processing and analyses utilizing R version 3.6.2. We compiled frequency tables for the overall prevalence of bloody diarrhea across age groups. We calculated odds ratios (OR) and confidence intervals (CI) in two distinct populations (bloody and non-bloody diarrheal cases) to assess the association between the presence of a pathogen and the likelihood of experiencing bloody diarrhea. In addition, we calculated OR adjusted by age group.

## Results

Among 1020 patients <18 years old with diarrhea, 126 (12%) of children or their caregivers reported bloody diarrhea (Supplemental Figure 2). PCR was performed on the stool samples of 111 (88%) cases of bloody diarrhea, of which 69 (62%) had at least one pathogen detected (Table 1 and Supplemental Figure 2). Among the 111 children with bloody diarrhea and PCR performed, we detected viruses in 20 (18%) of children, bacteria in 53 (48%), and protozoa (Giardia only) in 2 (2%). Two or more pathogens was detected in 22 children. Pathogens typically seen with bloody diarrhea, including *Campylobacter, C. difficile*, STEC, and *Shigella*, were collectively detected in 51 (46%). No pathogens were identified in 42 (38%) of children. In cases where only one pathogen was identified, we identified 32 (67%) with bacteria detected, 14 (29%) with viruses, and 2 (4%) with protozoa (Supplemental Table 3)

**Table 1:**
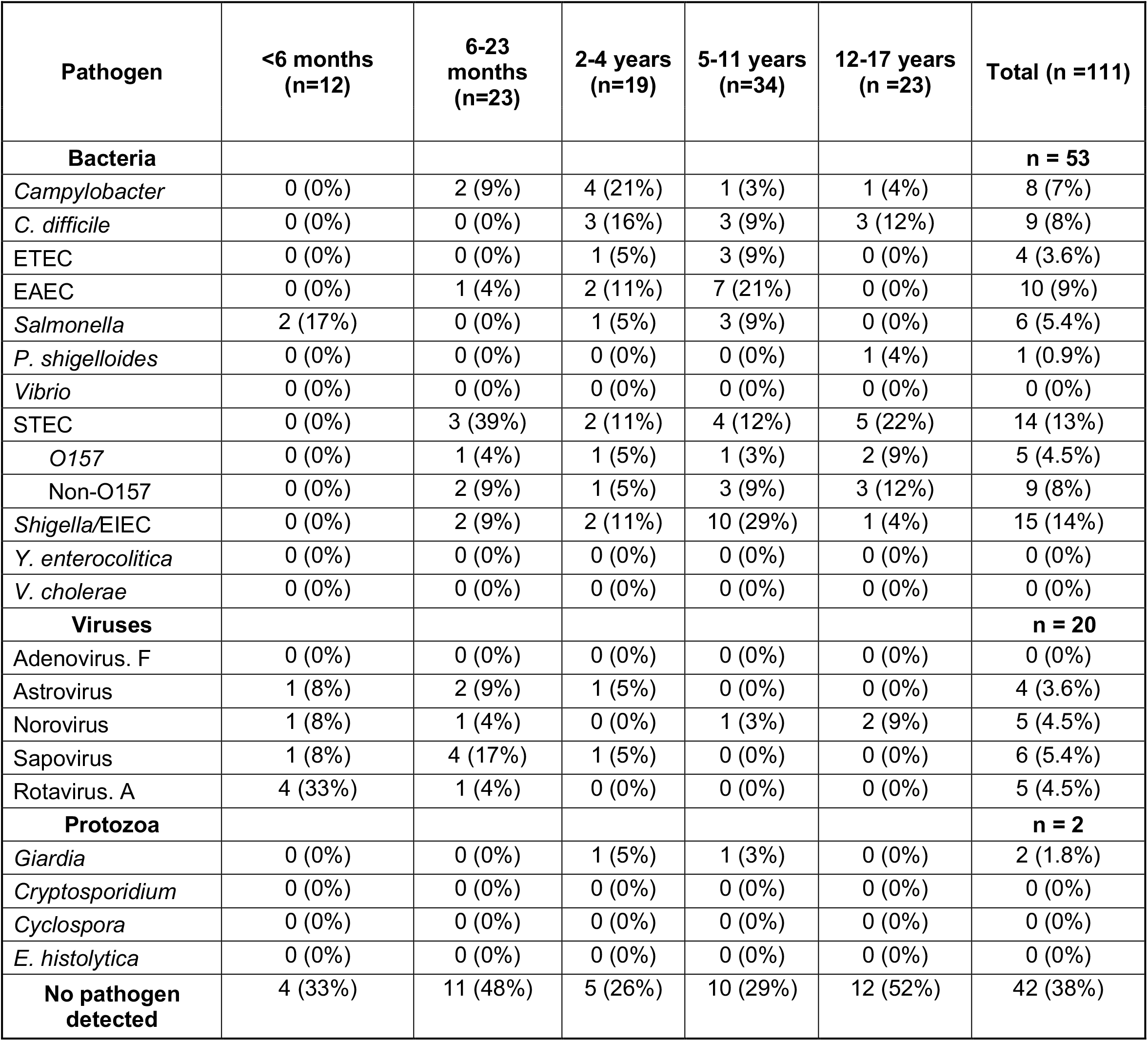
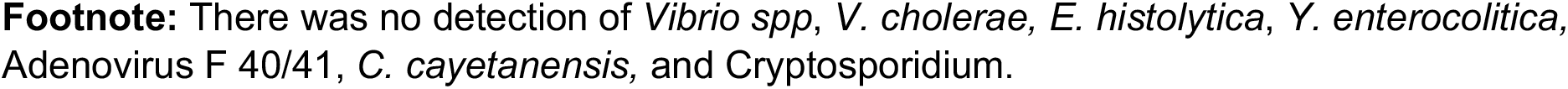
Frequency of detection for the 21 pathogens in the PCR panel, among patients presenting with bloody diarrhea, stratified by age group. Co-occurring pathogens are listed multiple times. Abbreviations: ETEC, Enterotoxigenic *E. coli*; EAEC, Enteroaggregative *E. coli*; STEC, Shigatoxin-producing *E. coli*; O157, *E. coli* O157

We conducted an odds ratio analysis to evaluate the magnitude of the association between the presence of pathogen and the occurrence of bloody diarrhea (Table 2), comparing patients with bloody diarrhea (n = 111), and those with non-bloody diarrhea (n=731). We found the odds of having bloody diarrhea are 9.4 times higher in individuals with STEC detected compared to those without (unadjusted OR: 9.4, 95% confidence interval [CI]: 4.1,21). After adjusting for age, the odds remain substantially elevated, with an adjusted OR of 8.9(95% CI:3.9,21). Similarly, for *Campylobacter*, the odds ratio was 4.3 (95% CI: 1.6,10.4). In contrast, sapovirus, astrovirus, and norovirus had lower odds of bloody diarrhea although the confidence interval excluded 1 only for norovirus and sapovirus. Of the 14 children with bloody diarrhea who received antibiotics during or after their ED visit (*Supplemental Table 4)*, 6 (43%) did not have a pathogen for which antibiotics would have been indicated; 3 patients had no pathogen detected, 1 had *Salmonella* detected in a non-infant, and 2 had STEC detected, for which antibiotics are potentially harmful.

**Table 2:**
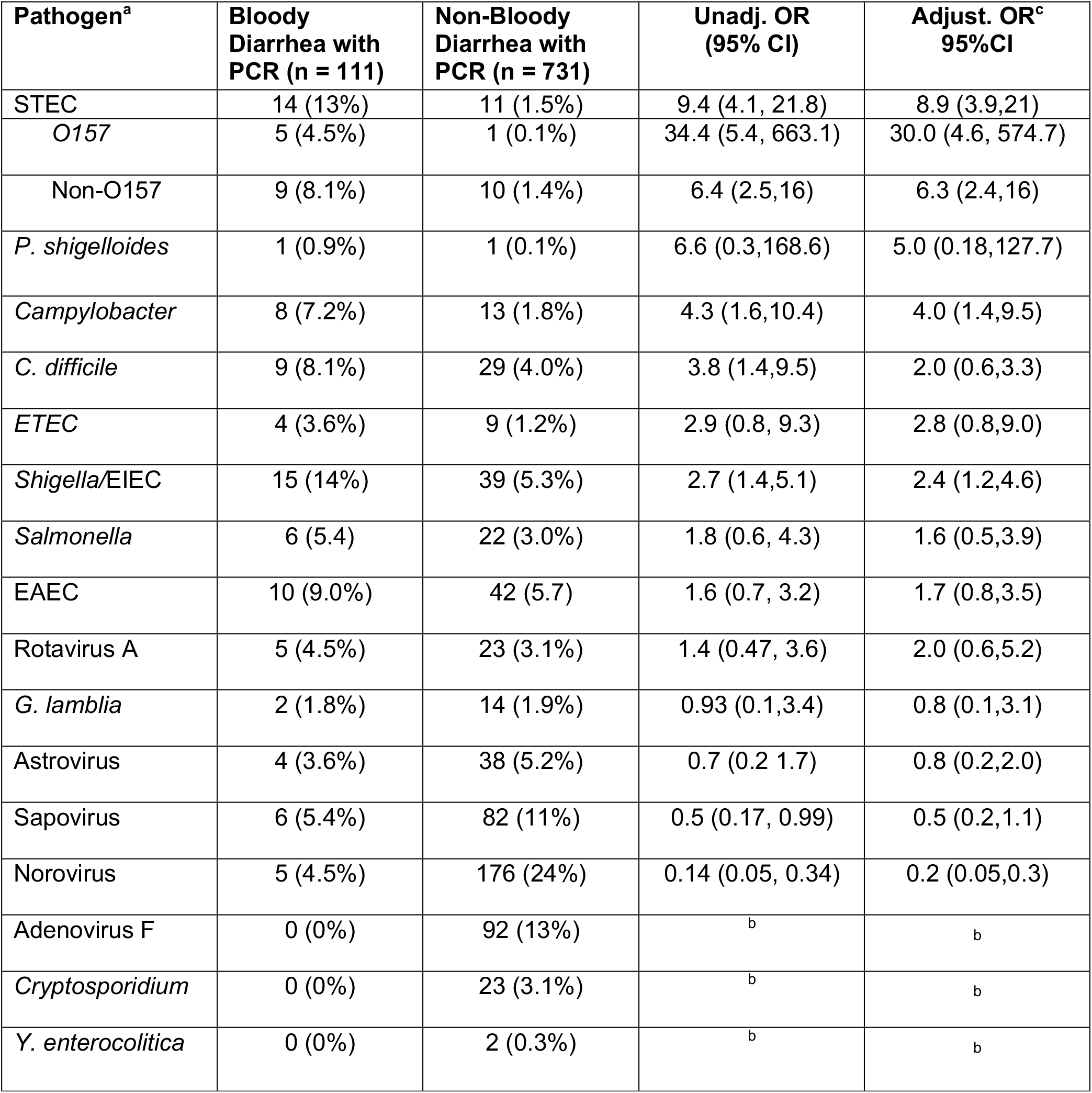
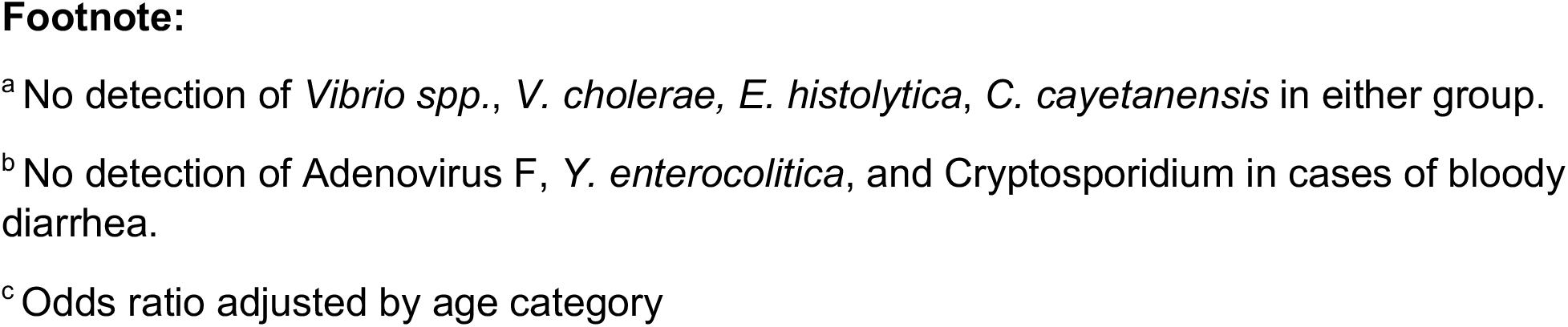
Unadjusted and age group-adjusted Odds Ratios (OR) of the association between the presence of pathogen and the occurrence of bloody diarrhea. Abbreviations: CI, confidence interval; OR odds ratio; Unadj, unadjusted; Adjust, adjusted.

## Discussion

Bloody diarrhea is thought to be a sign of invasive enteric infection that carries the risk of severe morbidity and mortality among children[4,10]. Our analysis of data from a multi-site study revealed that in less than half of the cases with bloody diarrhea was a bacterial pathogen typically associated with this condition detected, and that in a substantial proportion (over a third) of bloody diarrhea cases, a pathogen was detected for which antibiotics was not indicated. Prior studies of bloody diarrhea in children, before the widespread availability of multiplex PCR, had been limited to conventional stool culture, immunoassays, and a limited array of PCR detection methods[5,7,8]. Our findings align with more contemporary multiplex PCR-based studies of either bloody diarrhea, or more generally hematochezia, emphasizing the complexity of attributing bloody diarrhea symptoms to specific pathogens[4]. In our study, 13% (14/111) of children with bloody diarrhea had only viral detection, consistent with prior studies [4,11]. We also did not identify any pathogens in 38% of the patients, suggesting that a portion of bloody diarrhea cases could be attributed to non-infectious causes or some other pathogens causing gastrointestinal symptoms that the multiplex PCR panel may not cover. In addition, we found that 43% of patients with bloody diarrhea who received antibiotics during or after the ED visit did not have detection of a pathogen for which an antibiotic would be indicated. These findings support the need for use of stool testing in patients with bloody diarrhea to determine antibiotic use, instead of relying on empiric therapy.

While symptom-based empiric determination is insufficient to identify patients who would benefit from antibiotics, further work is also needed to improve the utility of diagnostic testing. Molecular testing such as multiplex PCR is an important improvement from traditional stool culture, with increased sensitivity for pathogen detection as well as faster turnaround time. However, molecular testing is expensive in high resource settings and may have very low availability in lower resource settings. Our data support its utility in bloody diarrhea, but more work is needed to identify strategies for optimal stewardship of this resource.

While our study provides valuable insights into the etiologies associated with bloody diarrhea in children presenting to emergency departments in the US, several limitations warrant consideration. The lack of control groups of non-diarrheal patients limits the ability to draw definitive conclusions about causation between identified pathogens and bloody diarrhea.

Second, in one of the five sites, a *Shigella* outbreak occurred during the study, which could lead to this pathogen being over-represented in our study. Third, our study only included patients presenting to emergency departments, and thus may represent more severe cases, restricting the generalizability of our findings. Despite these limitations, the study underscores the need for comprehensive testing and improved data collection strategies to improve antibiotic stewardship and patient care.

In conclusion, of the 12% (126/1020) of participants in this study of children presenting to US-based emergency rooms with acute diarrhea who reported bloody diarrhea, fewer than half of cases had etiologies for which antibiotics are indicated. Recognizing the heterogeneity of etiological agents in bloody diarrhea is crucial for the appropriate management of diarrheal illness in children.

## Data Availability

All data produced in the present study are available upon reasonable request to the authors.

## Acknowledgments

We thank all the field support staff who participated in this project. This study was partly funded by grants from the NIH (R01AI135114 and K24166087 to DTL). The IMPACT study was supported by the NIH with additional funding from BioFire Diagnostics (now bioMerieux).

This research was supported by the National Institutes of Health under Ruth L. Kirschstein National Research Service Award NIH Award Number 1T32TR00432

## References

1. Troeger C, Blacker BF, Khalil IA, et al. Estimates of the global, regional, and national morbidity, mortality, and aetiologies of diarrhoea in 195 countries: a systematic analysis for the Global Burden of Disease Study 2016. Published online 2018. doi:10.1016/S1473-3099(18)30362-1

2. UNICEF. Diarrhea remains a leading killer of young children, despite the availability of simple treatment solution. https://UNICEF.org/child-health/diarrhealdisease.html.

3. Scallan E, Mahon BE, Hoekstra RM, Griffin PM. Estimates of illnesses, hospitalizations and deaths caused by major bacterial enteric pathogens in young children in the United States. Pediatr Infect Dis J. 2013;32(3):217–221. doi:10.1097/INF.0B013E31827CA763

4. Böhrer M, Fitzpatrick E, Hurley K, et al. Hematochezia in children with acute diarrhea seeking emergency department care - a prospective cohort study. Acad Emerg Med. 2022;29(4):429–441. doi:10.1111/ACEM.14434

5. Xie J, Kim K, Berenger BM, et al. Comparison of a Rapid Multiplex Gastrointestinal Panel with Standard Laboratory Testing in the Management of Children with Hematochezia in a Pediatric Emergency Department: Randomized Controlled Trial. Microbiol Spectr. 2023;11(3). doi:10.1128/SPECTRUM.00268-23

6. Buchwald AG, Verani JR, Keita AM, et al. Etiology, Presentation, and Risk Factors for Diarrheal Syndromes in 3 Sub-Saharan African Countries After the Introduction of Rotavirus Vaccines From the Vaccine Impact on Diarrhea in Africa (VIDA) Study. Clinical Infectious Diseases. 2023;76(Supplement_1):S12–S22. doi:10.1093/CID/CIAD022

7. Buss SN, Leber A, Chapin K, et al. Multicenter Evaluation of the BioFire FilmArray Gastrointestinal Panel for Etiologic Diagnosis of Infectious Gastroenteritis. J Clin Microbiol. 2015;53(3):915. doi:10.1128/JCM.02674-14

8. Stockmann C, Pavia AT, Graham B, et al. Detection of 23 Gastrointestinal Pathogens Among Children Who Present With Diarrhea. J Pediatric Infect Dis Soc. 2017;6(3):231–238. doi:10.1093/JPIDS/PIW020

9. Talan DA, Moran GJ, Newdow M, et al. Etiology of Bloody Diarrhea among Patients Presenting to United States Emergency Departments: Prevalence of Escherichia coli O157:H7 and Other Enteropathogens. Clinical Infectious Diseases. 2001;573:573–580.

10. Denno DM, Shaikh N, Stapp JR, et al. Diarrhea Etiology in a Pediatric Emergency Department: A Case Control Study. Clin Infect Dis. 2012;55(7):897. doi:10.1093/CID/CIS553

11. Klein EJ, Boster DR, Stapp JR, et al. Diarrhea etiology in a Children’s Hospital Emergency Department: a prospective cohort study. Clin Infect Dis. 2006;43(7):807–813. doi:10.1086/507335

12. Shane AL, Mody RK, Crump JA, et al. 2017 Infectious Diseases Society of America Clinical Practice Guidelines for the Diagnosis and Management of Infectious Diarrhea. Clinical Infectious Diseases. 2017;65(12):e45–e80. doi:10.1093/cid/cix669

13. SB F, J X, MS N, et al. Shiga Toxin-Producing Escherichia coli Infection, Antibiotics, and Risk of Developing Hemolytic Uremic Syndrome: A Meta-analysis. Clin Infect Dis. 2016;62(10):1251–1258. doi:10.1093/CID/CIW099

14. Collins JP, King LM, Collier SA, et al. Antibiotic prescribing for acute gastroenteritis during ambulatory care visits-United States, 2006-2015. Infect Control Hosp Epidemiol. 2022;43(12):1880–1889. doi:10.1017/ICE.2021.522

15. Pavia AT, Cohen DM, Leber AL, et al. Clinical Impact of Multiplex Molecular Diagnostic Testing in Children with Acute Gastroenteritis Presenting to An Emergency Department: A Multicenter Prospective Study. medRxiv. Published online July 31, 2023:2023.07.27.23293208. doi:10.1101/2023.07.27.23293208

